# Feasibility, Safety, and Preliminary Efficacy of Partnered Rhythmic Rehabilitation in Prodromal Alzheimer’s Disease: A 12-Month Pilot Randomized Controlled Trial

**DOI:** 10.64898/2026.09.11.26362767

**Authors:** Forouzan Rafie, Daniel Azimi, Amir H. Nekouei, Haneul Kim, Cathleen M. Carroll-Sauer, Ihab Hajjar, Whitney Wharton, Deqiang Qiu, Felicia Goldstein, J. Lucas Mckay, Madeleine E. Hackney

**Affiliations:** Department of Medicine, Division of Geriatrics and Gerontology, Emory University School of Medicine, Atlanta, GA, United States, 30329; Emory University College of Arts and Sciences, Atlanta, GA, United States, 30322; Modeling in Health Research Center, Institute for Futures Studies in Health, Kerman University of Medical Sciences, Kerman, Iran; Oral and Dental Disease Research Center, Kerman University of Medical Sciences, Kerman, Iran; Department of Neurology, University of Texas Southwestern, Dallas, TX, United States, 75390; Department of Neurology, Baylor College of Medicine, TX, United States, 77030; Nell Hodgson Woodruff School of Nursing, Emory University, Atlanta, Georgia, United States, 30322; Department of Radiology and Imaging Sciences, Emory University School of Medicine, Atlanta, GA, United States, 30329; Department of Neurology, Emory University School of Medicine, Atlanta, GA, United States, 30322; Department of Biomedical Informatics, Emory University School of Medicine, Atlanta, GA, United States, 30322; Wallace H. Coulter, Department of Biomedical Engineering, Georgia Institute of Technology and Emory University, Atlanta, GA, United States, 30332; Goizueta Alzheimer’s Disease Research Center, Emory University, Atlanta, GA, United States, 30329; Department of Rehabilitation Medicine, Emory University School of Medicine, Atlanta, GA, United States, 30307; Atlanta VA Center for Visual & Neurocognitive Rehabilitation, Atlanta, GA, United States, 30319; Birmingham/Atlanta VA Geriatric Research Education and Clinical Center, Atlanta, GA, United States, 30319

**Author notes:** These authors contributed equally to this work (J. Lucas Mckay, Madeleine E. Hackney). Correspondence to:-Madeleine E. Hackney, Emory University, School of Medicine, Department of Medicine, Division of Geriatrics and Gerontology, Atlanta, GA 30329, USA., -Lucas McKay, Emory University, School of Medicine, Department of Biomedical Informatics, Atlanta, GA.

**Keywords:** Prodromal, Alzheimer’s disease, mild cognitive impairment, dance, walking, motor-cognitive integration, feasibility

## Abstract

**Background:** Non-pharmacological interventions may enhance motor-cognitive integration in prodromal Alzheimer’s disease (pAD). Partnered Rhythmic Rehabilitation (PRR), an adapted dance-based intervention, is designed to engage the motor, cognitive, and social domains. However, no randomized trial has evaluated its feasibility or efficacy in pAD. This study evaluated the feasibility, safety, and preliminary efficacy of PRR compared to group walking (WALK) in adults with pAD and amnestic mild cognitive impairment (aMCI).

**Methods:** In this single-blind, randomized controlled pilot trial (NCT04029623), 68 adults with pAD and aMCI were randomized to PRR or WALK. Prespecified feasibility criteria were attrition ≤15% and no injurious falls during interventions. Preliminary efficacy was examined on the primary outcome, the Four-Square Step Test (FSST). Secondary cognitive, motor, and psychosocial outcomes were also assessed to inform future definitive trials. Several modifications were made to the original protocol to ensure compliance with safety measures associated with the COVID-19 pandemic.

**Results:** Attrition in the PRR group (15.2%) did not significantly exceed the prespecified threshold (P = .848), and 0 injurious falls were reported during the supervised sessions, meeting both feasibility and safety criteria. No significant group-by-time interactions were observed for FSST, but significant group-by-time interactions favoring PRR were identified for Timed Up-and-Go (TUG; P = .021) and jump distance (P = .014).

**Conclusion:** PRR was feasible and safe in adults with aMCI consistent with pAD but did not meet the prespecified efficacy endpoint on FSST. Our results showed that TUG and standing board jump distance may serve as potential outcomes in future definitive trials.

## Introduction

Alzheimer’s disease (AD) is the most common neurodegenerative disorder, and the number of individuals living with dementia is projected to exceed 139 million by 2050 [1]. Prodromal Alzheimer’s disease (pAD), often characterized by mild cognitive impairment (MCI) attributable to AD, represents an important stage for intervention because annual progression rates to dementia are estimated at 10–15% [2]. In addition to cognitive decline, impairments in motor-cognitive integration may emerge early in the disease course and contribute to falls and functional dependence [3,4].

The increasing burden of AD reflects both population aging and the influence of modifiable risk factors, including physical inactivity and social isolation [5]. The 2024 Lancet Commission estimated that nearly half of all dementia cases could potentially be prevented or delayed through modification of known risk factors [6]. Although pharmacological treatments provide only modest symptomatic benefits, their effects on disease progression remain limited [7]. Consequently, there is substantial interest in non-pharmacological interventions that target mechanisms relevant to cognitive and functional decline.

Aerobic exercise promotes neuroprotection through reduced neuroinflammation and enhanced neurotrophic signaling [8,9]. Exercise interventions may engage motor, cognitive, and social domains, providing additive benefits through the recruitment of overlapping neural systems, including prefrontal and hippocampal networks [10]. While deficits in motor–cognitive integration are evident early in pAD [3], interventions targeting these domains simultaneously may be more efficacious for this population.

Dance-based intervention combines physical activity with cognitive and social engagement and therefore aligns with this multimodal approach. Dance requires continuous synchronization with rhythmic auditory cues, sensorimotor adaptation, and sustained attentional control, engaging auditory, cerebellar, and prefrontal networks [11]. Among individuals with Parkinson’s disease (PD), adapted Argentine tango has improved gait, balance, and postural control [12,13]. Given the overlap between motor and cognitive impairments observed in PD and pAD, these findings may have relevance for individuals with AD-related cognitive impairment [14]. Tango-based interventions have also been associated with improvement in attention, executive function, and spatial cognition in people with PD [15,16]. In addition, tango-based studies involving older adults with cognitive or mobility limitations have reported high levels of participation and retention [17]. Partnered dance may offer further advantages by incorporating interpersonal coordination, social interaction, and embodied cognitive processing [18,19].

To build on this evidence base, our group developed Partnered Rhythmic Rehabilitation (PRR), an adapted Argentine tango dance program designed to engage motor function, rhythmic entrainment, attention, working memory, and spatial cognition through continuous coordination with a dance partner [20]. Favorable adherence and satisfaction have also been reported in programs extending up to 16 months [21].

To our knowledge, no previous randomized controlled trial (RCT) has evaluated adapted tango in individuals with pAD [4]. This evidence gap is important because pAD presents unique clinical challenges, including impaired acquisition of new skills, increased fall risk, and reduced social participation. These characteristics may influence intervention feasibility, safety, adherence, and therapeutic response. Furthermore, the optimal duration, frequency, and delivery format of PRR for this population have not been established.

To address this gap, we conducted the PARTNER trial, a preregistered, single-blind, 12-month RCT (NCT04029623). The primary objective was to evaluate the feasibility, safety, and acceptability of PRR in adults with amnestic MCI consistent with pAD. A secondary objective was to examine the preliminary effects of PRR on motor–cognitive integration and functional mobility, with the Four-Square Step Test (FSST) designated as the primary efficacy outcome. Additional measures of motor-cognitive integration and functional mobility were included to provide a broader assessment of intervention effects and inform future definitive phase III trials. We hypothesized that PRR would demonstrate acceptable feasibility and safety, defined as attrition below the prespecified threshold of 15% and no injurious falls during supervised sessions. We further hypothesized that participants assigned to PRR would demonstrate greater improvement in FSST performance than control participants.

## Methods

### Study Design and Ethical Approval

PARTNER was a Phase II, 12-month, single-blind, parallel-arm RCT designed to evaluate the feasibility, safety, acceptability, and preliminary efficacy of PRR compared with walking (WALK) in individuals with pAD.

The study protocol was approved by the Emory University Institutional Review Board (IRB00110350, INSIGHT: 2024P007448). All participants provided written informed consent prior to their participation. Trial oversight was provided by a five-member Data Safety and Monitoring Board (DSMB) that met twice per year. This study was preregistered as the PARTNER trial (NCT04029623). Funding was provided by the National Institutes of Health, National Institute on Aging (NIH/NIA; 1R01AG062691-01).

### Study Population

Participants met criteria for aMCI consistent with pAD based on the Alzheimer’s Disease Neuroimaging Initiative (ADNI) diagnostic framework for aMCI [22,23], including subjective memory concern, objective memory impairment on standardized testing, global cognitive function within the MCI range, preserved functional independence, and absence of dementia.

All participants were aged 50 years or older. Recruitment was conducted through the Emory University Goizueta Alzheimer’s Disease Research Center using community outreach, collaboration with the Minority Engagement Core and Outreach and Recruitment Cores, the Emory Cognitive Empowerment Program, advertisements, health fairs, and research registries.

### Screening and Eligibility

Eligibility was determined with a multi-step process derived from ADNI study protocols as follows:

### Pre-screening

Telephone pre-screening was conducted by trained research staff to evaluate preliminary eligibility (i.e., evidence of pAD) and to minimize participant burden. Staff collected a brief medical history and administered the Montreal Cognitive Assessment, Blind/Telephone version (MoCA; cut-off score ≥ 18) [24]; the Telephone Interview for Cognitive Status (TICS; cut-off score ≥ 31) [25], the Functional Activities Questionnaire (FAQ; cut-off score ≤ 9) [26], and the Subjective Memory Concerns Questionnaire (SMQ; cut-off score ≥ 6) [27].

Individuals who met preliminary criteria were invited to complete a semi-structured Clinical Dementia Rating (CDR) interview [28]. Participants would be potentially eligible for the study if they had a Global CDR score of 0.5, consistent with questionable or mild cognitive impairment. The CDR is a validated 5-point scale assessing six domains of cognitive and functional performance; a Global CDR of 0.5 indicates early-stage cognitive decline without dementia.

### Screening

Written informed consent was obtained before any subsequent procedures. Participants completed a standardized assessment battery, including the Wechsler Memory Scale-Revised (WMS-R) Logical Memory delayed recall [29] and the full MoCA [24], with eligibility requiring a score between 18 and 25. Objective memory impairment was defined using education-adjusted ADNI cutoffs on the WMS-R Logical Memory delayed recall, with impairment defined as scores ≤11 (≥16 years of education), ≤9 (8–15 years), and ≤6 (≤7 years) [22,23]. The Edinburgh Handedness Inventory was also administered [30]. The neuropsychologist (FG) ultimately determined eligibility based on all pre-described clinical and neuropsychological criteria.

### Additional Inclusion and Exclusion Criteria

Additional inclusion criteria included the ability to walk at least 10 feet with or without an assistive device; currently performing less than 150 minutes per week of moderate-intensity physical activity or less than 75 minutes per week of vigorous-intensity physical activity; and no involvement in structured exercise programs during the previous three months.

One additional inclusion criterion was added after study initiation following consultation with the DSMB. After the first 11 participants were enrolled, the DSMB noted that some participants had not completed all baseline assessments prior to randomization. Consequently, completion of all baseline measures was added as an inclusion criterion.

Exclusion criteria encompassed medical, neurological, or psychiatric conditions that could compromise safe participation or confound study outcomes. These included acute illnesses requiring hospitalization, uncontrolled heart failure, a history of stroke within the previous three years, use of medications known to significantly affect cognition, and neurological disorders other than MCI (such as PD, multiple sclerosis, brain tumors, or seizure disorders), as well as major psychiatric illnesses.

### Randomization

Participants were randomized to PRR or WALK with 1:1 allocation, using the REDCap randomization module, stratified by age (≥65 or <65 years) and sex (male or female). Randomization lists were created by the study biostatistician in R software [31].

### Blinding

Participants were aware that they were enrolled in a rehabilitation intervention but were blinded to the study hypotheses. Due to resource limitations, instructors and outcome assessors were not blinded to group assignment. The study analysts and DSMB were blinded to group assignment until after all data collection was complete and a decision was made to break the randomization codes.

### Interventions

#### Session Frequencies

Both intervention arms followed a two-phase protocol of supervised 90-minute sessions. The Training Phase (months 1–3) comprised twice-weekly sessions over 12 weeks (24 possible sessions; minimum required to be considered adherent: 20). The Maintenance Phase (months 4– 12) comprised weekly sessions at a minimum of three per month for nine months (36 possible sessions; minimum required to be adherent: 27). Per-protocol intervention adherence was therefore defined as attendance at a minimum of 47 of 60 possible sessions (≥78%).

#### Partnered Rhythmic Rehabilitation (PRR)

PRR was delivered by trained instructors following a standardized curriculum. Each session consisted of a brief practice period of approximately 15 minutes to reinforce previously learned material, a 25-minute standing warm-up, and a 45-minute core training segment focused on partnered and rhythm-enhancing exercises, followed by a brief cool-down. During partnered activities, participants practiced both leading and following roles with trained study staff, students, volunteers, caregivers, or other support persons serving as dance partners. Partners facilitated safe practice, rhythmic and directional movement, weight transfer, and non-verbal communication of movement goals through touch. Partners rotated approximately every 15 minutes to allow participants to practice adapting to different movement partners. Instructors directed the curriculum and adapted task difficulty, while trained partners and staff monitored participants for fatigue, postural instability, and the need for rest or additional assistance. Heart rate and rating of perceived exertion were recorded five times during each session: upon arrival before exercise, immediately after the warm-up, and at 15-minute intervals during the 45-minute core training segment. These measurements were used to monitor exercise intensity and guide activity modification when needed [4].

#### Walking Intervention (WALK)

WALK was delivered in small groups by trained instructors following a standardized, manualized protocol. The intervention was matched to PRR in session frequency, duration, and target exercise intensity. Each session included a 25-minute standing warm-up that incorporated instruction and practice in safe walking mechanics, followed by a 45-minute structured walking segment with rest periods as needed and a brief cool-down. Participants were instructed to maintain an upright posture, use a natural stride length, and increase cadence when greater walking speed was appropriate.

Trained study staff, students, volunteers, caregivers, or other support people accompanied participants during the walking segment. These individuals provided social interaction comparable to that available in PRR, supported participant engagement and intervention delivery, and monitored participants for fatigue, gait instability, postural instability, and the need for rest or additional assistance. Unlike the dance partners in PRR, individuals accompanying WALK participants did not provide physical guidance or communicate movement goals through touch. The WALK instructor directed the session, reinforced safe walking mechanics, adjusted walking duration or intensity when needed, and retained responsibility for implementing safety procedures. Heart rate and rating of perceived exertion were recorded five times during each session: upon arrival before exercise, immediately after the warm-up, and at 15-minute intervals during the 45-minute walking segment. These measurements were used to monitor exercise intensity and guide activity modification when needed [4].

#### Prespecified Outcomes

We prespecified outcomes related to attrition, adherence, safety, satisfaction, and preliminary efficacy as follows:

#### Attrition

Attrition was defined as withdrawal from the study before completion of the intervention period for any reason. Attendance was tracked continuously by research staff, and missed sessions prompted follow-up contact to identify and address barriers to participation. Attrition was prespecified as a feasibility criterion, with attrition ≤15% considered acceptable; this threshold was consistent with feasibility benchmarks used in early-phase behavioral intervention trials [32]. Observed attrition was compared against this threshold using a one-sided binomial test.

#### Intervention Adherence

Adherence was quantified as the number of intervention sessions each participant attended. Per-protocol dose was defined as attending ≥47 intervention sessions (nominally ≥20 during the Training Phase and ≥27 during the Maintenance Phase) over the nominal one-year observation period. Attendance was recorded by staff and entered into REDCap.

#### Safety

Safety was defined as the occurrence of injurious falls attributable to intervention participation and was monitored continuously during all supervised sessions. Trained staff prospectively monitored and documented adverse events, including falls, near-falls, injuries, and medical complaints occurring during or immediately after sessions. Any adverse event triggered clinical review and, when appropriate, referral for medical evaluation. The primary safety endpoint was the incidence of injurious falls during study participation. We prespecified that the intervention would be considered safe if no injurious falls attributable to intervention participation occurred during supervised sessions.

#### Participant Satisfaction

Participant satisfaction was assessed at the conclusion of the intervention using a structured exit questionnaire that included items on program enjoyment, perceived physical and mental activity, enhanced knowledge or skills, influence on self-care, willingness to attend future programs, and willingness to continue participating if the program remained available. Responses were recorded on a 5-point Likert scale ranging from strongly disagree to strongly agree. For descriptive reporting, responses of agree or strongly agree were classified as favorable.

#### Outcome Assessments

The primary efficacy endpoint was change in motor-cognitive integration over time, quantified using FSST performance across repeated assessments in both groups.

#### Clinical and Demographic Variables

Clinical and demographic characteristics, including age, sex, fall history, and other relevant health and background factors, were obtained through structured participant self-report.

#### Primary Outcome: The Four-Square Step Test (FSST)

FSST was the prespecified primary efficacy outcome measure and was used to assess motor– cognitive integration through rapid stepping, movement sequencing, change of direction, and coordination [33]. FSST has been associated with executive function and dual-task mobility demands in older adults and neurological populations [34]. Participants stepped in clockwise and counterclockwise sequences through four squares formed by rods arranged in a cross configuration on the floor. They were instructed to complete the sequence as quickly and safely as possible without touching the rods. Trials were timed using a stopwatch. If a participant touched or displaced a rod, the trial was stopped and repeated. Participants completed trials until three valid attempts were obtained, and the fastest valid completion time was used for analysis. Lower scores indicated better performance.

### Secondary Outcomes

#### Motor-Cognitive and Motor Outcomes

Secondary outcomes assessed motor-cognitive integration, balance, postural control, gait, functional mobility, lower-extremity strength and power, aerobic endurance, and turning performance. Motor-cognitive integration was assessed with the Timed Up-and-Go (TUG) test under single-task, manual dual-task, and cognitive dual-task conditions [35–37]. The manual dual-task condition required participants to carry a cup of water, and the cognitive dual-task condition required serial subtraction by threes. The Body Position Spatial Test (BPST) [38] was also a test of motor-cognitive integration.

Balance and postural control were assessed with the Mini-Balance Evaluation Systems Test (Mini-BESTest) [39], Dynamic Gait Index (DGI) [40], tandem walk interruptions [41], tandem stance time [42], and one-leg stance time [42]. Tandem stance and one-leg stance values were averaged across left and right limbs.

Gait speed and cadence were assessed during preferred forward walking, fast forward walking, and backward walking over a 20-foot (6.1-m) walkway. Three trials were completed for each condition, and mean values across trials were used for analysis [43]. Lower-extremity strength was assessed with the 30-second Chair Stand Test [44], and lower-extremity functional power was assessed using standing broad jump distance [45]. Aerobic endurance was assessed with the Six-Minute Walk Test (6MWT) [46,47]. Turning performance was assessed with the 360° Turn Test, recorded as the average number of steps and time required to complete left and right turns [48]. For time-based outcomes, lower values indicated better performance; for distance, repetition, balance, and gait speed outcomes, higher values indicated better performance.

#### Cognitive Outcomes

Global cognition was assessed with the MoCA [24] . Executive function and cognitive flexibility were assessed with Trail Making Test (TMT) Parts A and B and the TMT B–A difference score [49]. Response inhibition and set-shifting were assessed with the Color–Word Interference Test (CWIT) from the Delis–Kaplan Executive Function System [50]. Planning and psychomotor speed were assessed using Tower of London (TOL) indices, including the total achievement score, mean first-move time, and the time-per-move ratio [51]. Cognitive dual-task performance was assessed as serial subtraction accuracy during the cognitive TUG condition [37]. Verbal learning and memory were assessed with the Rey Auditory Verbal Learning Test (RAVLT), including immediate recall, long-delay free recall, recognition correct responses, and recognition false positives [52]. Visuospatial memory was assessed with the Rey Complex Figure Test (RCFT), including immediate recall time, delayed recall score, and copy time [52,53]. Working memory was assessed with Digit Span Forward and Backward [54], the Brooks Spatial Memory task [55], the Reverse Corsi Block Test, and the Corsi Blocks product score [56]. Visuospatial processing was assessed with the Benton Judgment of Line Orientation test (BJLO) [57], and language was assessed with the Boston Naming Test (BNT) [58].

#### Psychosocial and Self-Reported Surveys

Psychosocial and patient-reported outcomes assessed depressive symptoms, health-related quality of life, functional independence, balanced confidence, physical activity, community mobility, participation, autonomy, and perceived social support. Depressive symptoms were assessed using the Beck Depression Inventory-II (BDI-II) [59], the Center for Epidemiologic Studies Depression Scale (CES-D) [60] and the Patient Health Questionnaire-9 (PHQ-9) [61]. Health-related quality of life was assessed with the Quality of Life in Alzheimer’s Disease scale (QoL-AD) [62] and the 12-Item Short-Form Health Survey (SF-12) with physical composite and mental composite scales (PCS/MCS) [63]. Functional independence was assessed with the Instrumental Activities of Daily Living scale (IADL) [64].

Balance confidence was assessed with the Activities-specific Balance Confidence scale (ABC) [65]. Physical activity was assessed with the Paffenbarger Physical Activity Questionnaire and the Physical Activity Scale for the Elderly (PASE) [66,67]. Community mobility was assessed with the Life Space Questionnaire [68]. Participation and autonomy were assessed with the Impact on Participation and Autonomy questionnaire (IPA) [69]. Perceived social support was assessed with the Multidimensional Scale of Perceived Social Support (MSPSS) [70].

#### Prespecified Statistical Analysis

Descriptive statistics were generated separately for each study arm and summarized as means and standard deviations for continuous variables and as counts and percentages for categorical variables. Attrition was examined as a feasibility endpoint using a one-sided comparison against the prespecified null proportion of 15%. Intervention adherence was examined using chi-square analysis.

Statistical analysis was conducted on an intent-to-treat basis for all randomized participants using R (version 4.5.1) [31] in the RStudio environment. Analysis of Fall History and Intervention Adherence was conducted in Excel (version 16.110.3, Microsoft Corporation). The primary efficacy endpoint, FSST performance, expressed in seconds, with lower values indicating superior performance, was assessed at baseline, 3 months, and 12 months in both the PRR and WALK groups. Time was dummy coded with baseline as the reference level, and WALK served as the reference group for the group variable. The FSST score and secondary outcomes were analyzed using a linear mixed-effects model with fixed effects for group, time, and their interaction in the *lmerTest* (version 3.2.1) package. Fall history was examined using chi-square analysis. An alpha level of 0.05 was used for all hypothesis tests. Exit survey responses were summarized descriptively by intervention group using item-level frequencies and response distribution to demonstrate the distributions between the PRR and WALK groups.

#### Protocol Changes to Accommodate the COVID-19 Pandemic

Because the trial was conducted during the COVID-19 pandemic between February 2021 and September 2024, numerous changes were required after study initiation to ensure participant safety. Following review by the DSMB in January 2021, the following temporary changes were made and implemented until January 2023, when restrictions eased:

• The enrollment period was delayed to begin after the availability of the COVID-19 vaccine (January 2021) to study staff and participants and extended to accommodate reduced throughput.

• Electronic consent for participants was implemented. Consent forms were emailed in advance for review, reviewed with the participants during remote screening visits, and signed through a REDCap link.

• Sessions were delivered in a hybrid format for some participants who were unable or unwilling to come to in-person visits. Within each block of five sessions, the first was conducted in person, and the remaining ones were delivered virtually via video conference for both arms. Participants could attend additional in-person visits. Personal Protective Equipment (PPE) was used consistently by study staff and participants. In addition to face coverings, which were provided by the team, if necessary, in place of the modified embrace frame used by participants in previous PRR interventions, each pair of participants held opposite ends of a commercially available toy “hula hoop” to ensure adequate separation.

## Results

### Recruitment

A schematic diagram depicting participant flow through the study is presented in Figure 1. A total of 199 individuals were assessed for eligibility by telephone screen. After exclusions, 68 participants were randomized to either PRR (n = 33) or WALK (n = 35). Enrollment occurred from October 2019 through June 2024, with follow-up concluding in September 2025. The trial ended after all enrolled participants completed the planned 12-month assessment. The COVID-19 pandemic prolonged the duration of the study, but the study proceeded to completion as planned.

**Figure 1.**
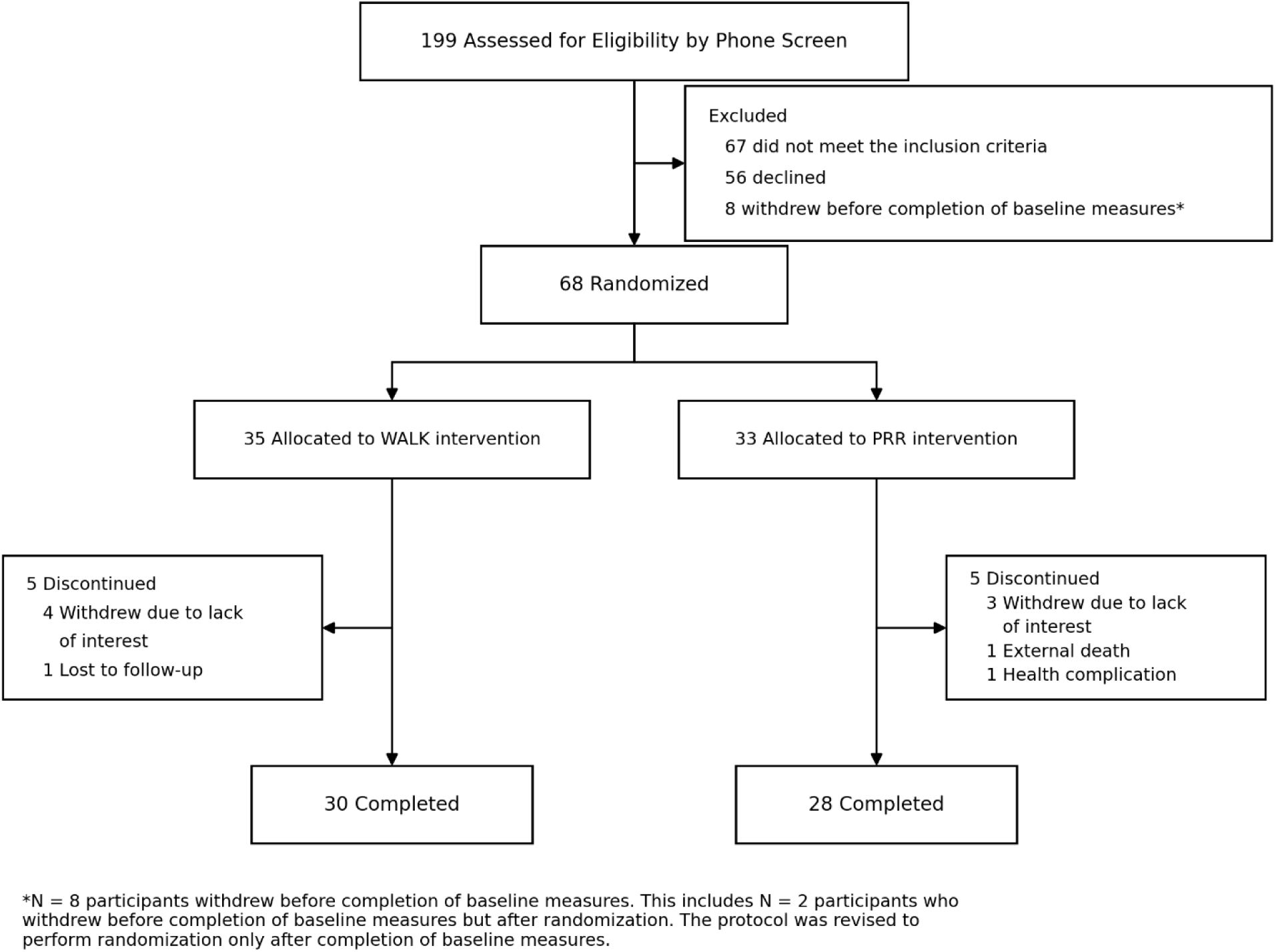
Consort Diagram

### Participant Characteristics

Demographic and clinical characteristics of the sample are presented in Table 1. The mean age across both intervention arms was 74.6 ± 7.1, and 30/68 (44%) participants were male.

**Table 1.** Demographic and clinical characteristics of participants, stratified by intervention arm.

|  | **PRR**<br>**(N=33)** | **WALK**<br>**(N=35)** | **Total**<br>**(N=68)** |
| --- | --- | --- | --- |
| **Age (years)** | 75.1 (6.0) | 74.1 (8.0) | 74.6 (7.1) |
| **Sex** |  |  |  |
| Female | 19 (58%) | 19 (54%) | 38 (56%) |
| Male | 14 (42%) | 16 (46%) | 30 (44%) |
| **Education (years)** | 15.6 (2.5) | 16.1 (2.3) | 15.9 (2.4) |
| **Race/Ethnicity** |  |  |  |
| White/Caucasian | 17 (52%) | 19 (54%) | 36 (53%) |
| Black/African American | 13 (39%) | 14 (40%) | 27 (40%) |
| Multiracial | 3 (9%) | 0 (0%) | 3 (4%) |
| Hispanic or Latino | 0 (0%) | 2 (6%) | 2 (3%) |
| **BMI** | 28.4 (4.0) | 26.8 (5.8) | 27.5 (5.1) |
| **Marital Status** |  |  |  |
| Married/Partner | 20 (61%) | 23 (66%) | 43 (63%) |
| Separate/Divorced | 4 (12%) | 7 (20%) | 11 (16%) |
| Widowed | 5 (15%) | 5 (14%) | 10 (15%) |
| Single | 4 (12%) | 0 (0%) | 4 (6%) |
| **Housing** |  |  |  |
| House/apartment/condominium | 28 (85%) | 32 (91%) | 60 (88%) |
| Senior housing (independent) | 3 (9%) | 1 (3%) | 4 (6%) |
| Assisted living | 0 (0%) | 2 (6%) | 2 (3%) |
| Relative's home | 2 (6%) | 0 (0%) | 2 (3%) |
| **Occupational Status** |  |  |  |
| Retired | 30 (91%) | 29 (83%) | 59 (87%) |
| Work part-time | 2 (6%) | 2 (6%) | 4 (6%) |
| Disabled | 1 (3%) | 1 (3%) | 2 (3%) |
| Homemaker | 0 (0%) | 2 (6%) | 2 (3%) |
| Work full-time | 0 (0%) | 1 (3%) | 1 (1%) |
| **Number of Comorbidities** | 4.3 (2.2) | 3.6 (2.0) | 3.9 (2.1) |
| **Fall History (Past year)** |  |  |  |
| No Falls | 19 (58%) | 28 (80%) | 47 (69%) |
| ≥1 Fall | 14 (42%) | 7 (20%) | 21 (31%) |
| **Assisted Device Use** |  |  |  |
| No | 28 (85%) | 30 (86%) | 58 (85%) |
| Sometimes | 2 (6%) | 3 (9%) | 5 (7%) |
| Yes | 3 (9%) | 2 (6%) | 5 (7%) |
| **Quality of Life<sup>1</sup>** | 6.2 (0.9) | 5.9 (1.1)<sup>a</sup> | 6.0 (1.0)<sup>b</sup> |
| **BDI-II** | 6.9 (5.4) | 5.8 (4.4) | 6.3 (4.9) |
| **MoCA** | 23.0 (3.7) | 24.2 (3.7) | 23.6 (3.7) |
| **FAQ** | 2.8 (3.8) | 1.6 (2.2) | 2.2 (3.1) |
| **TICS** | 30.3 (3.6) | 31.9 (2.4) | 31.1 (3.1) |
| **CPF** | 21.3 (3.6) | 22.2 (3.0)<sup>a</sup> | 21.8 (3.3)<sup>b</sup> |

**Abbreviations:** BMI, Body Mass Index; BDI-II, Beck Depression Inventory-II; FAQ, Functional Activities Questionnaire; TICS, Telephone Interview for Cognitive Status; CPF, Composite Physical Function Values are presented as n (%) for categorical variables and mean (SD) for continuous variables.
<sup>1</sup> Quality of Life: 1=very low, 7=very high.
<sup>a</sup> N=34; <sup>b</sup> N=67.

### Attrition

Ten participants withdrew prior to study completion (PRR: n = 5, 15.2%; WALK: n = 5, 14.3%). Fifty-eight participants completed the 12-month assessment (PRR, n = 28; WALK, n = 30). Attrition did not significantly exceed the prespecified 15% feasibility threshold in either arm (one-sided P = .848) (Table 2).

**Table 2.**
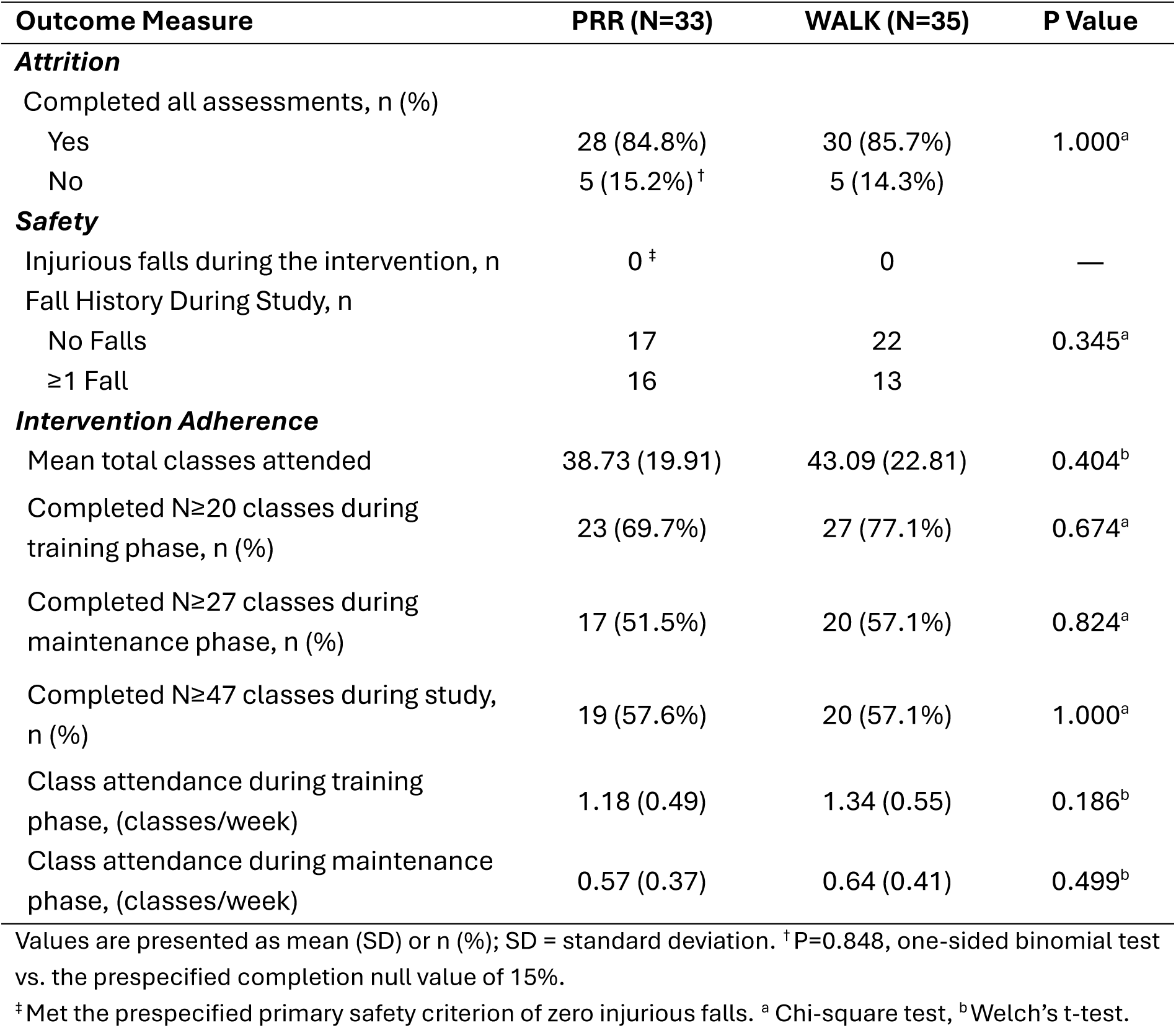
Feasibility and safety outcomes stratified by intervention arm.

### Intervention Adherence

The average number of completed intervention sessions was 38.7 ± 19.9 in PRR and 43.1 ± 22.8 in WALK (P = .404, t-test). Per protocol attendance was 19/33 (57.6%) participants in PRR and 20/35 (57.1%) participants in WALK (P = 1.000, chi-squared test; Table 2).

### Safety

No adverse events, including injurious falls, near-falls, or medical complaints attributable to intervention or assessment participation, were reported during approximately 1200 supervised PRR sessions or across WALK sessions throughout the 12-month study period, confirming the prespecified safety criteria for both arms (Table 2).

### Participant Satisfaction

Of the 58 participants who completed the 12-month intervention, 55 responded to the Exit Questionnaire (PRR: n = 27; WALK: n = 28). Among PRR respondents, 24 agreed or strongly agreed that they enjoyed participating in the program, 21 reported being more physically active, 25 reported being more mentally active, 26 indicated that the program enhanced their knowledge or skills, 25 reported that the program would influence how they take care of themselves, 24 indicated that they would attend future programs, and 23 indicated that they would continue participating if possible. Responses to the remaining questionnaire items were also favorable. The complete item-level response distributions for both groups are presented in Figure 2.

**Figure 2.**
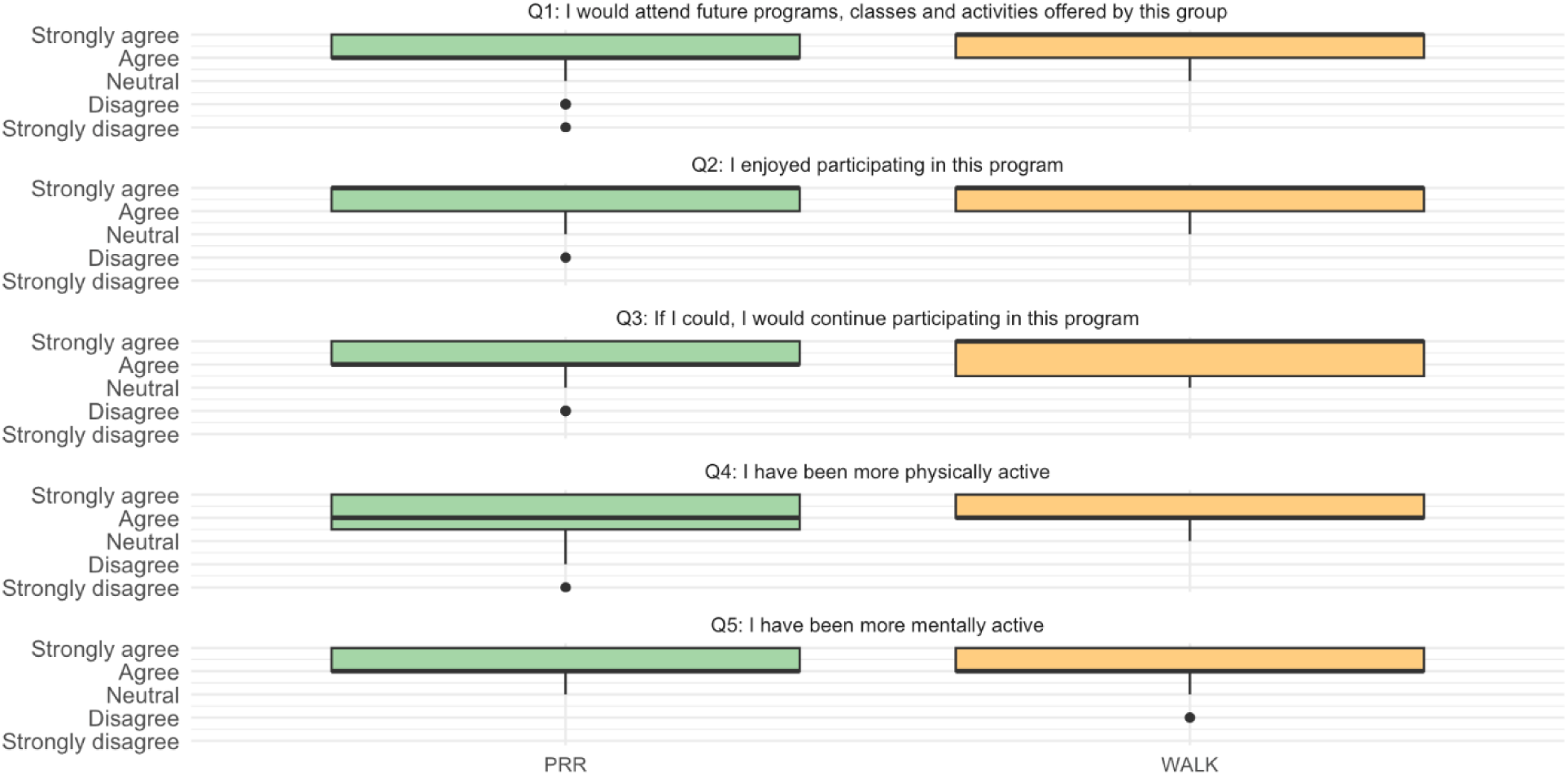
Distribution of participant satisfaction scores from the exit survey across PRR (N=27) and WALK (N=28) groups. The boxes represent the interquartile range, and the dots represent outliers.

### Preliminary Efficacy

#### Primary Outcome: The Four-Square Step Test (FSST)

Participants in both arms showed non-significant improvement in the primary outcome, FSST performance, at 3 months; PRR FSST completion time changed by –0.46 seconds and WALK FSST completion time changing by –0.50 seconds from baseline to 3 months, and the group × time interaction was not significant at 3 months (P = .645). WALK retained a modest improvement at 12 months relative to baseline, changing by +0.13 seconds from 3 months to 12 months for an overall change of –0.37 seconds from baseline until the end of the study. PRR rose above baseline at 12 months, changing +1.12 seconds from 3 months to 12 months for an overall change of +0.66 seconds from baseline until the end of the study; the group × time interaction at 12 months did not reach significance (P = .053; Table 3).

**Table 3.** Motor outcome measures stratified by intervention arm and timepoint.

| Outcome Measure | Time Point | PRR (n=33) | WALK (n = 35) | P Value |
| --- | --- | --- | --- | --- |
| ***Balance and Postural Control*** |  |  |  |  |
| Four-square step test (FSST), s | Baseline | 12.66 (3.76) | 12.31 (3.76) | — |
|  | 3 months | 12.20 (3.14) | 11.81 (3.23) | 0.645 |
|  | 12 months | 13.32 (4.81) | 11.94 (2.81) | 0.053 |
| Body Position Spatial Test-Span (BPST), score (0-9) | Baseline | 3.24 (0.71) | 3.11 (0.83) | — |
|  | 3 months | 3.21 (0.82) | 3.57 (1.01) | 0.080 |
|  | 12 months | 3.21 (0.69) | 3.34 (0.86) | 0.344 |
| Mini-BESTest, Total Score (0-28) | Baseline | 23.39 (3.20) | 22.94 (3.08) | — |
|  | 3 months | 22.31 (3.30) | 23.80 (2.33) | 0.002* |
|  | 12 months | 22.39 (3.86) | 23.48 (2.34) | 0.020^ |
| Dynamic Gait Index (DGI), total score (0-24)<sup>b</sup> | Baseline | 21.85 (2.46) | 22.26 (1.82) | — |
|  | 3 months | 21.97 (1.95) | 22.63 (1.38) | 0.588 |
|  | 12 months | 21.96 (2.16) | 22.28 (1.83) | 0.877 |
| Tandem Walk Interruptions, n | Baseline | 3.45 (3.11) | 3.00 (2.91) | — |
|  | 3 months | 4.11 (2.88) | 3.00 (2.63) | 0.332 |
|  | 12 months | 3.78 (3.58) | 3.52 (3.53) | 0.887 |
| Tandem Stance time (average left/right), s | Baseline | 21.82 (9.86) | 23.33 (8.35) | — |
|  | 3 months | 22.46 (10.08) | 22.83 (8.71) | 0.617 |
|  | 12 months | 22.87 (10.13) | 22.69 (9.41) | 0.248 |
| One-Leg Stance (average left/right), s | Baseline | 13.14 (10.14) | 12.37 (9.48) | — |
|  | 3 months | 12.45 (9.84) | 11.91 (9.40) | 0.582 |
|  | 12 months | 13.16 (9.88) | 10.94 (9.39) | 0.613 |
| ***Gait Speed and Cadence*** |  |  |  |  |
| Comfortable Gait Speed, m/s | Baseline | 1.00 (0.22) | 0.99 (0.20) | — |
|  | 3 months | 1.03 (0.25) | 1.07 (0.21) | 0.300 |
|  | 12 months | 1.01 (0.29) | 1.02 (0.19) | 0.716 |
| Comfortable Gait Cadence, steps/min | Baseline | 108.75 (14.72) | 104.69 (15.94) | — |
|  | 3 months | 110.15 (17.98) | 111.24 (13.30) | 0.151 |
|  | 12 months | 106.85 (17.65) | 107.28 (11.91) | 0.173 |
| Fast Gait Speed, m/s | Baseline | 1.43 (0.35) | 1.47 (0.34) | — |
|  | 3 months | 1.43 (0.34) | 1.51 (0.35) | 0.262 |
|  | 12 months | 1.30 (0.45) | 1.41 (0.32) | 0.129 |
| Fast Gait Cadence, steps/min | Baseline | 129.73 (17.74) | 130.89 (21.67) | — |
|  | 3 months | 132.36 (19.03) | 137.85 (22.32) | 0.350 |
|  | 12 months | 120.11 (30.24) | 128.82 (19.45) | 0.151 |
| Backward Gait Speed, m/s | Baseline | 0.63 (0.24) | 0.63 (0.25) | — |
|  | 3 months | 0.63 (0.25) | 0.68 (0.25) | 0.235 |
|  | 12 months | 0.59 (0.23) | 0.64 (0.21) | 0.301 |
| Backward Gait Cadence, steps/min | Baseline | 112.41 (20.56) | 103.08 (21.19) | — |
|  | 3 months | 107.70 (21.92) | 111.89 (19.63) | 0.004* |
|  | 12 months | 102.59 (19.20) | 102.78 (17.29) | 0.046^ |
| **Functional Mobility** |  |  |  |  |
| TUG – Simple condition, s | Baseline | 10.89 (2.88) | 9.22 (2.05) | — |
|  | 3 months | 10.23 (2.54) | 9.33 (2.55) | 0.251 |
|  | 12 months | 10.16 (2.57) | 9.90 (2.50) | 0.021^ |
| TUG – Manual condition, s | Baseline | 14.03 (3.92) | 12.59 (3.02) | — |
|  | 3 months | 13.83 (4.21) | 12.28 (3.16) | 0.555 |
|  | 12 months | 13.81 (3.52) | 12.64 (3.00) | 0.907 |
| TUG – Cognitive condition, s | Baseline | 17.88 (6.19) | 17.06 (7.77) | — |
|  | 3 months | 17.52 (6.94) | 16.04 (9.22) | 0.814 |
|  | 12 months | 19.62 (13.40) | 17.11 (8.67) | 0.502 |
| **Strength and Power** |  |  |  |  |
| 30s Chair Stand test, repetitions | Baseline | 10.34 (3.23) | 11.40 (3.48) | — |
|  | 3 months | 10.68 (2.63) | 11.50 (3.34) | 0.955 |
|  | 12 months | 11.00 (3.56) | 11.63 (3.43) | 0.458 |
| Standing broad jump distance, m | Baseline | 0.47 (0.34) | 0.63 (0.38) | — |
|  | 3 months | 0.64 (0.44) | 0.63 (0.27) | 0.088 |
|  | 12 months | 0.79 (0.50) | 0.65 (0.32) | 0.014^ |
| **Aerobic capacity and endurance** |  |  |  |  |
| Six-minute walk test distance (6MWT), m<sup>b</sup> | Baseline | 372.59 (89.87) | 392.78 (118.13) | — |
|  | 3 months | 357.78 (87.37) | 429.01 (128.40) | 0.028* |
|  | 12 months | 372.75 (100.10) | 385.18 (108.02) | 0.802 |
| **Turning Performance** |  |  |  |  |
| 360° turn steps, n (average left/right) | Baseline | 6.98 (1.62) | 6.67 (1.10) | — |
|  | 3 months | 7.47 (2.15) | 7.00 (1.46) | 0.508 |
|  | 12 months | 7.16 (2.38) | 7.15 (1.65) | 0.511 |
| 360° turn time, s (average left/right) | Baseline | 3.54 (1.40) | 3.38 (1.29) | — |
|  | 3 months | 3.62 (1.51) | 3.44 (1.39) | 0.540 |
|  | 12 months | 3.55 (1.41) | 3.61 (1.37) | 0.961 |

**Abbreviations:** PRR, Partnered Rhythmic Rehabilitation; WALK, walking exercise; FSST, Four Square Step Test; TUG, Timed Up-and-Go; DGI, Dynamic Gait Index; Mini-BESTest, Mini Balance Evaluation Systems Test; BPST, Body Position Spatial Test; 6MWT, 6-Minute Walk Test.
Gait speed and cadence measures represent averages across left and right trials. Tandem stance and one-leg stance values represent averages across three trials per limb.
**Score direction:** Higher scores indicate better performance for: Mini-BESTest, BPST, DGI, 30-second chair stand (repetitions), gait speed (all conditions), cadence (all conditions), tandem stance, one leg stance, jump distance, and 6MWT distance. Lower scores indicate better performance for: FSST, all TUG variants, 360° turn time and step count, and tandem walk interruptions.
Values are presented as mean (SD); SD = standard deviation; all P values represent between-group differences in changes from baseline to 3 and 12 months (linear mixed model).
\* Statistically significant ( $P < 0.05$ ) group × time interaction, baseline to 3 months; ^Statistically significant ( $P < 0.05$ ) group × time interaction, baseline to 12 months.

#### Secondary Outcomes

Several secondary outcomes showed significant group × time interactions. The simple TUG condition demonstrated a significant group × time interaction at 12 months (P = 0.021). The standing broad jump distance showed a significant group × time interaction at 12 months, supporting the PRR (P = 0.014). Mini-BESTest scores were significantly higher in WALK at both 3 months (P = 0.002) and 12 months (P = 0.020). Backward walking cadence showed significant group differences in WALK at 3 months (P = 0.004) and 12 months (P = 0.046). The 6MWT distance was significantly greater in WALK at 3 months (P = 0.028), though this difference was not present by 12 months (P = 0.802). Serial subtraction accuracy during the cognitive dual-task condition was significantly higher in WALK at 3 months (P = 0.013), with no difference at 12 months (P = 0.472). No significant group differences were observed for any cognitive outcome (Table 4) or psychosocial measure (Table 5).

**Table 4.**
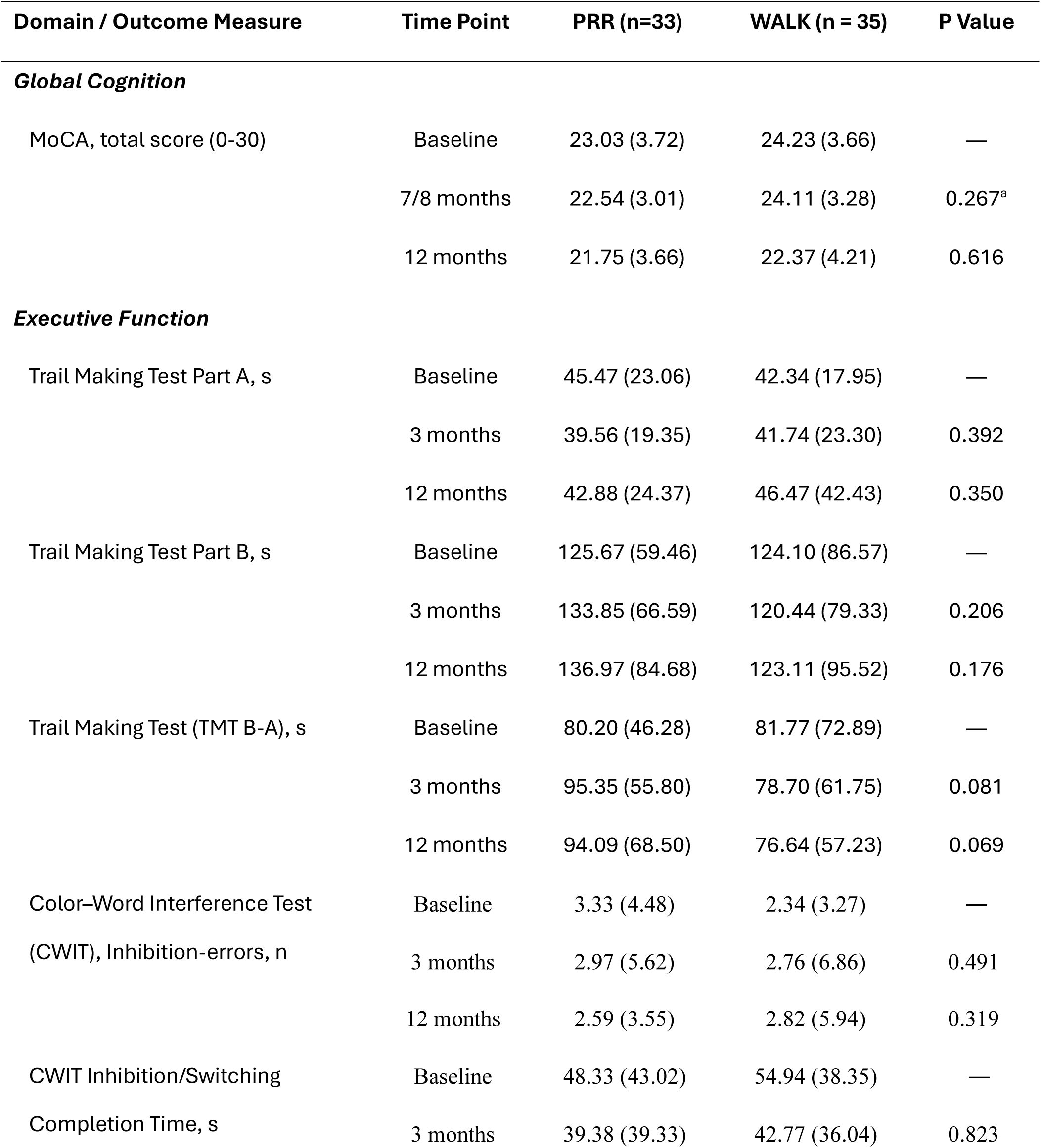
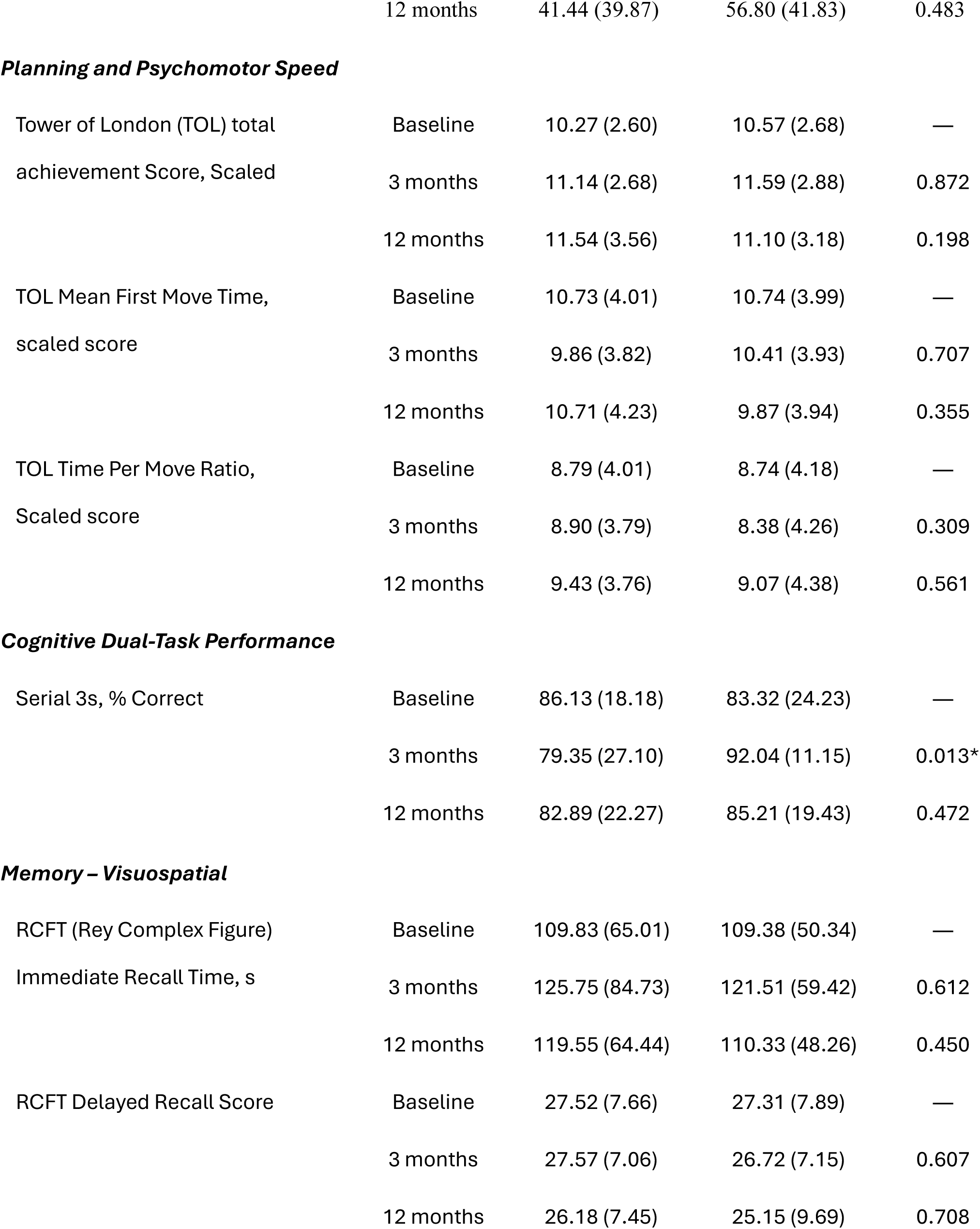
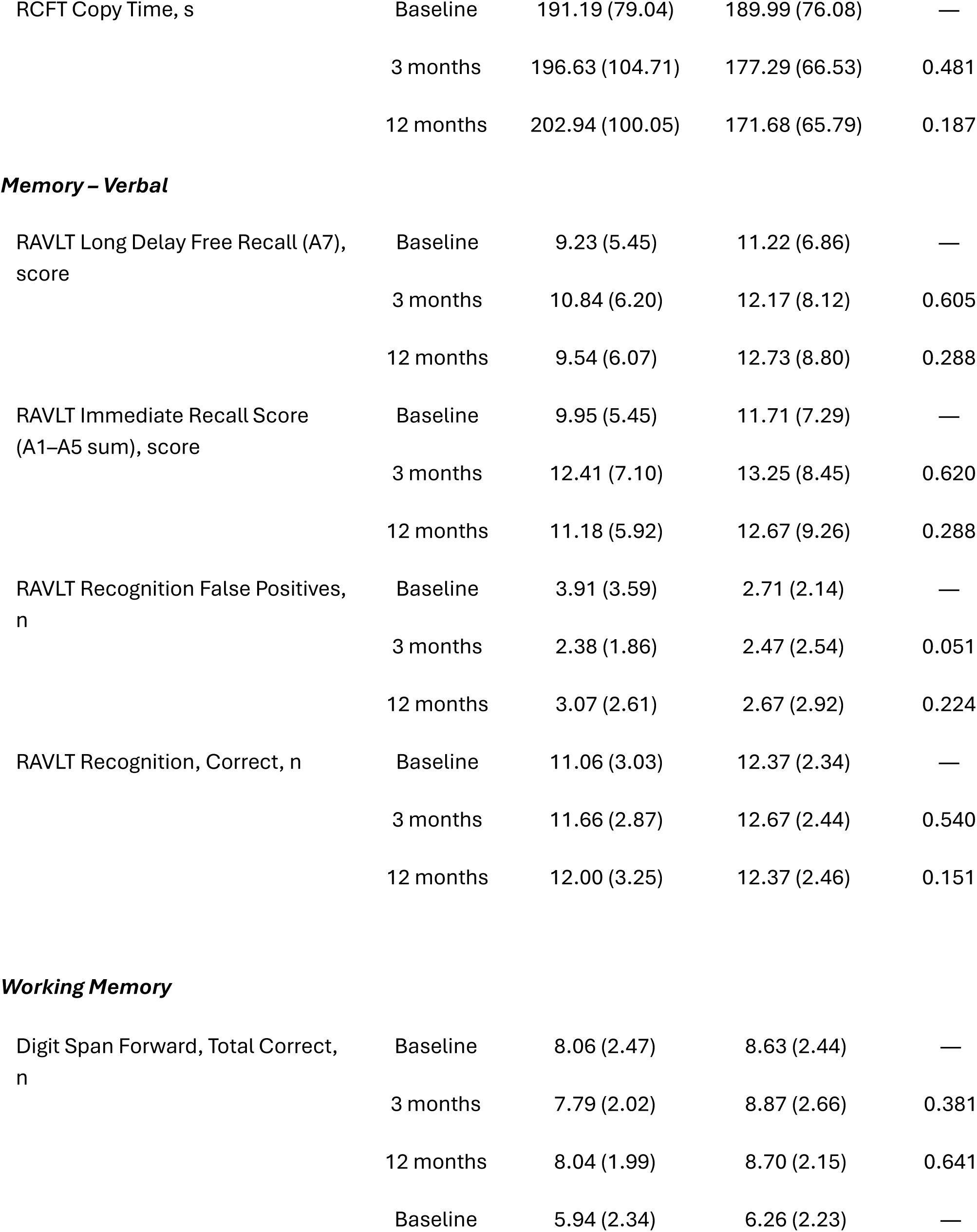
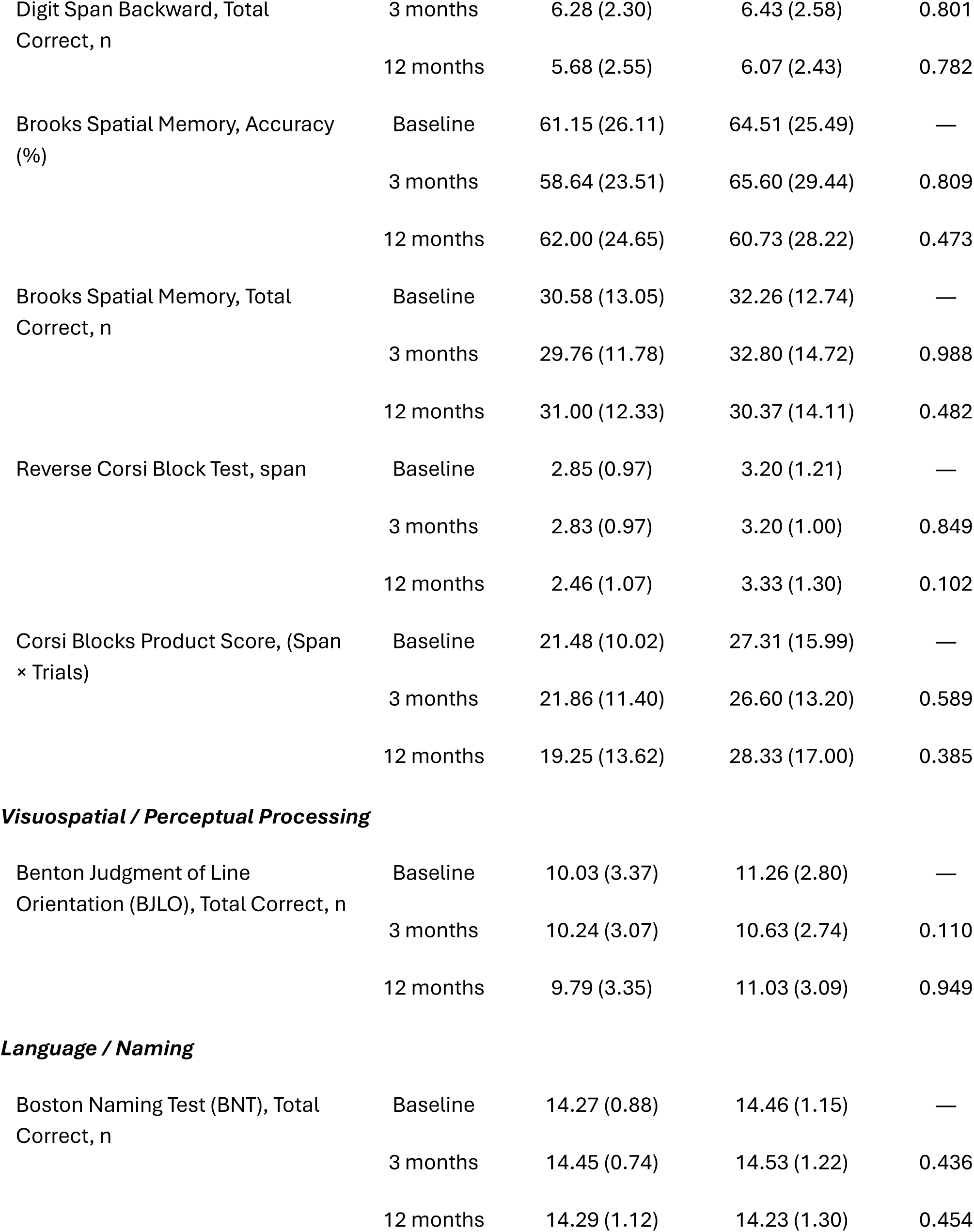
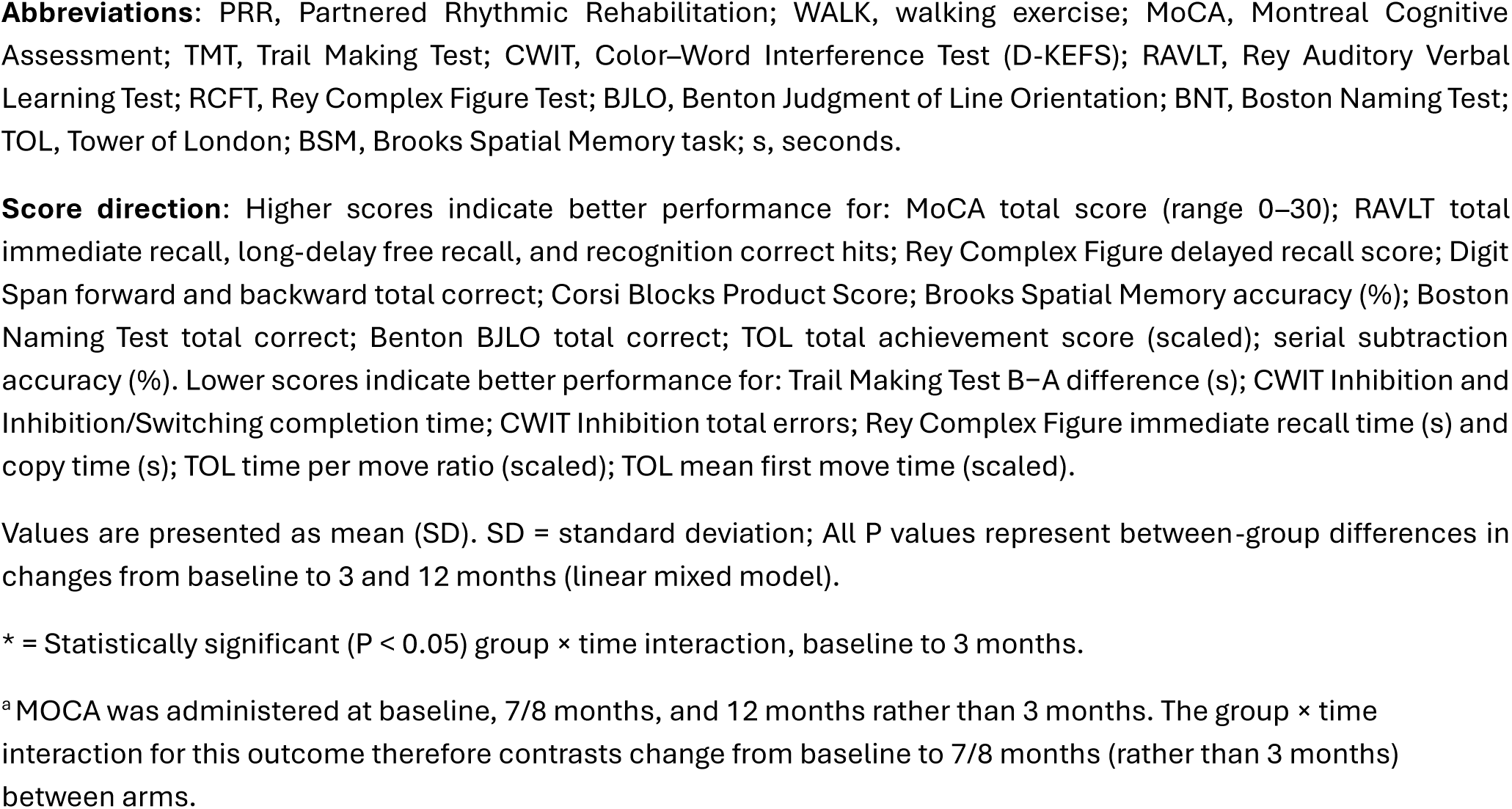
Cognitive outcome measures stratified by intervention arm and timepoint.

**Table 5.** Psychosocial outcome measures stratified by intervention arm and timepoint.

| Domain / Outcome Measure | Time Point | PRR (n=33) | WALK (n = 35) | P Value |
| --- | --- | --- | --- | --- |
| ***Depressive & Psychological Symptoms*** |  |  |  |  |
| Beck Depression Inventory (BDI-II), Total score | Baseline | 6.88 (5.41) | 5.80 (4.42) | — |
|  | 3 months | 6.43 (5.38) | 5.28 (6.00) | 0.860 |
|  | 12 months | 6.68 (6.36) | 5.67 (4.41) | 0.827 |
| Center for Epidemiologic Studies Depression Scale (CES-D), total score | Baseline | 7.09 (6.36) | 7.59 (7.84) | — |
|  | 3 months | 7.00 (6.24) | 6.55 (7.62) | 0.527 |
|  | 12 months | 8.68 (7.29) | 6.79 (6.88) | 0.071 |
| Patient Health Questionnaire-9 (PHQ-9) total score | Baseline | 3.42 (3.46) | 3.06 (3.60) | — |
|  | 3 months | 3.00 (2.85) | 3.04 (3.86) | 0.519 |
|  | 12 months | 3.29 (3.31) | 2.90 (2.34) | 0.995 |
| ***Health-Related Quality of Life*** |  |  |  |  |
| Quality of Life in Alzheimer's Disease (QoL AD), total score | Baseline | 40.39 (5.36) | 40.38 (5.53) | — |
|  | 3 months | 40.61 (4.93) | 40.17 (5.82) | 0.624 |
|  | 12 months | 41.46 (5.59) | 40.83 (5.89) | 0.496 |
| SF-12 Physical Component Summary (PCS), component score | Baseline | 50.77 (10.02) | 50.97 (9.63) | — |
|  | 3 months | 50.55 (8.37) | 52.33 (9.07) | 0.196 |
|  | 12 months | 49.41 (8.56) | 52.77 (8.94) | 0.088 |
| SF-12 Mental Component Summary (MCS), component score | Baseline | 45.40 (5.99) | 44.65 (5.92) | — |
|  | 3 months | 44.71 (5.00) | 45.84 (4.65) | 0.217 |
|  | 12 months | 44.47 (5.02) | 45.88 (4.82) | 0.162 |
| ***Functional Independence & Physical Function*** |  |  |  |  |
| Instrumental Activities of Daily Living (IADL) score, 0–8 | Baseline | 6.44 (1.74) | 7.18 (1.33) | — |
|  | 3 months | 6.67 (2.06) | 7.45 (1.13) | 0.951 |
|  | 12 months | 5.75 (2.43) | 7.55 (1.92) | 0.154 |
| ***Balance Confidence*** |  |  |  |  |
| Activities-specific Balance Confidence scale (ABC), score | Baseline | 85.85 (13.37) | 89.32 (14.09) | — |
|  | 3 months | 88.06 (12.37) | 90.04 (12.93) | 0.404 |
|  | 12 months | 87.39 (10.06) | 89.77 (10.79) | 0.604 |
| ***Physical Activity & Community Mobility*** |  |  |  |  |
| Paffenbarger Physical Activity Total, kcal/week | Baseline | 2568.06 (2962.01) | 4154.49 (6616.13) | — |
|  | 3 months | 2384.24 (3195.86) | 3351.77 (3417.42) | 0.500 |
|  | 12 months | 2161.76 (3437.35) | 2103.84 (2500.09) | 0.124 |
| Physical Activity Scale for the Elderly (PASE), total score | Baseline | 124.91 (71.80) | 129.31 (84.10) | — |
|  | 3 months | 109.13 (68.74) | 137.52 (71.01) | 0.270 |
|  | 12 months | 106.47 (71.99) | 123.89 (64.43) | 0.853 |
| ***Community Mobility*** |  |  |  |  |
| Life Space Questionnaire | Baseline | 10.91 (0.54) | 11.17 (1.03) | — |
| Total | 3 months | 11.09 (1.30) | 11.08 (1.24) | 0.555 |
|  | 12 months | 11.10 (1.66) | 11.33 (1.15) | 0.958 |
| **Participation, Autonomy** |  |  |  |  |
| Impact on Participation and | Baseline | 17.64 (12.87) | 15.66 (15.93) | — |
| Autonomy (IPA), total score | 3 months | 15.82 (13.47) | 14.03 (17.26) | 0.579 |
|  | 12 months | 17.11 (14.59) | 14.83 (14.90) | 0.833 |
| **Social Support** |  |  |  |  |
| Multidimensional Scale of | Baseline | 5.90 (1.10) | 5.96 (1.23) | — |
| Perceived Social Support | 3 months | 6.33 (0.82) | 6.11 (0.75) | 0.600 |
| (MSPSS), total score | 12 months | 5.60 (1.89) | 6.09 (0.96) | 0.185 |

**Abbreviations:** PRR, Partnered Rhythmic Rehabilitation; WALK, walking exercise; IADL, Instrumental Activities of Daily Living; BDI, Beck Depression Inventory; SF-12, 12-Item Short-Form Health Survey; PCS, Physical Component Summary; MCS, Mental Component Summary; IPA, Impact on Participation and Autonomy; ABC, Activities-specific Balance Confidence scale; CES-D, Center for Epidemiologic Studies Depression Scale; MSPSS, Multidimensional Scale of Perceived Social Support; PHQ-9, Patient Health Questionnaire-9; QoL-AD, Quality of Life–Alzheimer's Disease; PASE, Physical Activity Scale for the Elderly. **Score direction:** Higher scores indicate better outcomes for QoL-AD total score; SF-12 PCS and MCS component scores (norm-based, population mean = 50, SD = 10); IADL score (range 0–8); ABC scale score (range 0–100%); Paffenbarger Physical Activity Total (kcal/week); PASE total score; Life Space Questionnaire total score; MSPSS total score (range 1–7). Lower scores indicate better outcomes for BDI total score; CES-D total score; PHQ-9 total score; and IPA total score.
Values are presented as mean (SD). SD = standard deviation; All P values represent between-group differences in changes from baseline to 3 and 12 months (group × time interaction from linear mixed-effects models).
\* = Statistically significant ( $P < 0.05$ ) group × time interaction, baseline to 3 months.

Within executive function, the number of errors in CWIT for PRR decreased over time, while the number of errors in WALK increased (3.33 to 2.59 and 2.34 to 2.82 from baseline to 12 months, respectively). The number of RAVLT recognition false positives and correct answers improved in PRR; the number of false positives decreased over time in PRR (3.91 to 3.07) while relatively remaining steady in WALK (2.71 to 2.67), and the number of correct answers increased over time in PRR (11.06 to 12.00) while remaining the same in WALK (12.37 to 12.37). The 3-month group × time interaction for RAVLT recognition false positives was near significant (P = .051). Finally, Brooks spatial memory performance among PRR participants was maintained over time (61.15% to 62.00%), while performance worsened among WALK participants (64.51% to 60.73%) (Table 3).

##### Additional analyses

The proportion of participants reporting at least one fall during the study did not differ significantly between groups (PRR: n = 16 with ≥1 fall; WALK: n = 13; P = 0.345, Chi-squared test, *post hoc*) (Table 2).

## Discussion

The PARTNER trial evaluated the feasibility, safety, acceptability, and preliminary efficacy of a 12-month PRR program in adults with clinically defined amnestic MCI consistent with pAD. PRR met the prespecified attrition and safety criteria, and satisfaction was high among participants who completed the exit survey. The prespecified intervention dose of at least 47 sessions was completed by 58% of participants assigned to PRR. Among PRR participants who initiated the intervention but did not reach this threshold, the mean attendance was 23.92 ± 15.33 sessions (range, 1–46), corresponding to approximately 51% of the intended dose. Adherence was similar between groups. No injurious falls attributable to the intervention occurred during supervised sessions, supporting the safety of supervised PRR delivery within the context of this pilot trial. PRR did not demonstrate superiority over WALK on the prespecified primary outcome, the FSST. However, several exploratory secondary outcomes favored PRR, suggesting that partnered dance may target specific domains of mobility and motor-cognitive integration not fully captured by the primary endpoint.

The proportion of participants completing the study in PARTNER was broadly consistent with prior exercise and dance-based studies involving older adults with cognitive impairment. The BAILAMOS trial reported 42.8% attrition over 16 weeks among community-dwelling older Latino adults with MCI [71], the 12-month MOVE for your MIND pilot trial reported intervention adherence rates of 56%–62% among older adults with MCI or mild dementia [72], and the GERAS DANCE study reported 20% attrition over 15 weeks among older adults with early cognitive and mobility impairments [73]. In PARTNER, approximately 85% of randomized participants completed the 12-month assessment despite pandemic-related disruptions, although achieving the intended intervention dose remained challenging.

The absence of intervention-attributable injurious falls during supervised sessions was consistent with prior dance-based feasibility studies. Bennett et al. reported no falls during 12 weeks of adapted dance sessions in individuals with AD and related dementias [74], whereas the GERAS DANCE study reported one non-injurious fall during supervised sessions [73]. These findings support the safety of appropriately adapted and supervised dance-based interventions over the 12 months in this population.

Participants in both intervention groups who completed the exit survey reported high satisfaction. PRR respondents provided favorable feedback regarding program enjoyment, mental activity, enhanced knowledge or skills, and willingness to continue participating. WALK respondents reported higher frequencies of favorable responses for program enjoyment and perceived physical activity, although these item-level patterns were not formally compared between groups. Favorable responses across both intervention groups are consistent with Kim et al. (2026), who found that partnered dance and structured walking may support different dimensions of program engagement in people with PD [21]. These findings support the acceptability of long-term PRR among participants who remained engaged through follow-up.

PRR did not produce greater improvement than WALK on the prespecified primary FSST outcome. Both groups improved during the 3-month training phase, but these changes were not sustained at 12 months. The lower prescribed frequency during the maintenance phase may have contributed to this pattern, although the trial was not designed to establish a dose-response relationship. More frequent assessments will be needed to characterize the timing and durability of changes in FSST performance.

Among the exploratory secondary motor outcomes, group-by-time interactions favoring PRR were observed for simple TUG and standing broad jump distance, PRR requires movement initiation, controlled turning, anticipatory postural adjustment, directional transitions, and coordinated weight transfer, demands that overlap with the motor sequencing and lower-extremity power requirements of both outcomes. The progressive improvement in TUG performance in PRR, contrasted with slowing in WALK, represents a potentially PRR-specific pattern of mobility change, although the mean improvement did not exceed the minimal detectable change (MDC90) of 4.09 s derived from individuals with established AD [75]. Because that threshold was developed for individual-level interpretation in a more impaired population, the clinical significance of this finding in adults with amnestic MCI remains uncertain. Standing broad jump distance also favored PRR, suggesting a potential training effect on lower-extremity explosive power, although the mean improvement did not exceed the published MDC90 [76].

WALK showed advantages in Mini-BESTest performance and an early advantage in 6MWT distance. Mean changes in Mini-BESTest within both groups did not reach the MDC95 of 3.5 points or the 4-point threshold for clinically meaningful improvement in adults with balance disorders [77]. The mean 3-month improvement in 6MWT distance in WALK exceeded the MDC of 33.47 m reported in individuals with AD [75] and the 20-m threshold proposed for a small meaningful change in older adults [78], although this improvement was not significant at 12 months. Because these thresholds were not established specifically in adults with amnestic MCI, these comparisons require cautious interpretation.

Serial subtraction accuracy during the cognitive dual-task condition favored WALK at 3 months, but this difference did not remain significant at 12 months. No other cognitive outcome demonstrated a significant group-by-time interaction, although the direction of effects for selected outcomes, including CWIT, RAVLT, and Brooks, favored PRR. In particular, the 3-month group-by-time effect size for RAVLT false positives was small to moderate (d = −0.43) and favored PRR, suggesting that this measure may warrant consideration in future PRR trials.

A key strength of the PARTNER trial was its rigorous, multistep participant screening and selection process, which increased confidence that participants represented the clinical pAD phenotype.

## Limitations

This study had several limitations. A substantial portion of the trial was conducted during the COVID-19 pandemic, and social-distancing requirements likely reduced adherence below anticipated levels. Whether these modifications had differential effects across arms could not be formally evaluated, although WALK did not require physical partnering and may have been more amenable to remote delivery. The lower prescribed frequency during the maintenance phase may have affected the durability of intervention-related changes, although the trial was not designed to establish a dose-response relationship. However, the maintenance dose was associated with largely retained performance on several motor and cognitive measures by participants.

## Conclusion

The PARTNER trial showed that a 12-month PRR program was feasible, safe when delivered under supervision, and acceptable to older adults with amnestic MCI consistent with pAD, although adherence to the intended intervention dose was lower than planned. PRR did not improve FSST performance relative to WALK, although simple TUG and standing broad jump distance had significant interactions and will inform outcome selection for future trials.

## Data Availability

All data produced in the present study are available upon reasonable request to the authors

## Acknowledgments

The authors thank Sherman A. Jones for assistance with data organization. We also thank all students and research staff who supported study operations and the participants and their families whose time and commitment made this research possible.

## Author Contributions

Madeleine E. Hackney conceptualized and designed the study and acquired funding. Ihab Hajjar, Whitney Wharton, and Felicia Goldstein supported study design. Forouzan Rafie, Cathleen M. Carroll-Sauer, and Haneul Kim participated in data collection and curation. J. Lucas Mckay and Daniel Azimi conducted statistical analysis, interpreted the data, and provided critical comments and input on research output. Amir H. Nekouei conducted analysis and visualization. Forouzan Rafie wrote the original draft. Madeleine E. Hackney, J. Lucas Mckay, Forouzan Rafie, and Daniel Azimi provided critical revisions to the manuscript. All authors read the manuscript and approved the final version for publication.

## Data Availability Statement

The data supporting the findings of this study are available on request from the corresponding author.

